# SMOOTH protocol: A pilot randomised prospective intra-patient single-blinded observational study for examining the mechanistic basis of ablative fractional carbon dioxide laser therapy in treating hypertrophic scarring

**DOI:** 10.1101/2023.04.19.23288792

**Authors:** Yung-Yi Chen, Krupali M. Patel, Rizwana Imran, Tarek Hassouna, Ezekwe Amirize, Abdulrazak Abdulsalam, Jonathan Bishop, Anita Slade, Maximina Ventura, Jeremy Yarrow, Janet M. Lord, Yvonne Wilson, Naiem S. Moiemen

## Abstract

**Background:** Burn injuries are the fourth most common type of trauma and are associated with substantial morbidity and mortality. The impact of burn injury is clinically significant as burn injuries often give rise to exuberant scarring. Hypertrophic scarring (HTS) is a particular concern as up to 70% of burns patients develop HTS. Laser therapy is used for treating HTS and has shown positive clinical outcomes, although the mechanisms remain unclear limiting approaches to improve its effectiveness. Emerging evidence has shown that fibroblasts and senescent cells are important modifiers of scarring. This study aims to investigate the cellular kinetics in HTS after laser therapy, with a focus on the association of scar reduction with the presence of senescent cells.

**Methods:** We will conduct a multicentre, intra-patient, single-blinded, randomised controlled longitudinal pilot study with parallel assignments to achieve this objective. 60 participants will be recruited to receive 3 interventional ablative fractional CO_2_ laser treatments over a 12-month period. Each participant will have two scars randomly allocated to receive either laser treatment or standard care. Biopsies will be obtained from laser-treated, scarred- no treatment and non-scarred tissues for immune-histological staining to investigate the longitudinal kinetics of p16^INK4A+^-senescent cells and fibroblast subpopulations (CD90^+^/Thy1^+^ and αSMA^+^). Combined subjective scar assessments including Modified Vancouver Scar Scale, Patient and Observer Scar Assessment Scale and Brisbane Burn Scar Impact Profile; and objective assessment tools including 3D-Vectra-H1 photography, DermaScan^®^ Cortex, Cutometer^®^ and ColoriMeter^®^DSMIII will be used to evaluate clinical outcomes. These will then be used to investigate the association between senescent cells and scar reduction after laser therapy. This study will also collect blood samples to explore the systemic biomarkers associated with the response to laser therapy.

**Discussion:** This study will provide an improved understanding of mechanisms potentially mediating scar reduction with laser treatment, which will enable better designs of laser treatment regimens for those living with HTS.

**Trial registration:** ClinicalTrials.gov: NCT04736251.

## Introduction

Burn injury is the fourth most common type of trauma after road traffic accidents, falls and interpersonal intentional injury and is associated with substantial morbidity and mortality. According to the World Health Organisation, burn injuries account for around 180,000 deaths worldwide annually, with approximately 11 million people requiring medical attention each year.(1) The cost of wound care in clinical practice over 24 months in the UK is estimated around £16,924 per burn.(2) The impact of burn injuries is clinically significant as burn injuries often give rise to exuberant scarring that results in permanent physical function loss and psychological issues due to the stigma of disfigurement. These outcomes ultimately lead to profound long-term effects on quality of life (QoL). Up to 90% of the patients who survive a burn injury suffer from post-burn scarring.(3, 4) Among different types of scarring, hypertrophic scarring is a particular concern in deep, full and partial thickness burn injuries and up to 70% of patients develops hypertrophic scars (HTS) following burns.(5)

Burn care during the last 30 years has seen a step change in survival and this has been paralleled by improved acute care and durable wound cover resulting in less deformity and scarring. However, there remains an urgent need for improvements in post-burn scar assessment, management and the treatment of historic scars. Gangemi et al in a comprehensive review of 703 burn survivors’ records identified the key risk factors for post-burn hypertrophic scarring.(6) These factors included age, gender, dark skin, burn severity, number of surgical procedures performed to achieve wound cover, burn location (neck and/or upper limbs) and time to wound healing. How these factors influence the treatment of established scars remains poorly understood.

### Treatments for post-burn HTS

Post-burn HTS is typically treated non-invasively with the use of topical emollients, silicone gel, compression garments or these modalities in various combinations. A survey of 19 paediatric burn services in the UK showed that 18 services routinely use pressure garments for prevention of HTS following burn injury.(7) More recently, intra-lesional therapy using steroids or anti-neoplastic drugs such as fluorouracil, bleomycin and interferon have come into use.(8, 9) Other commonly used drugs include verapamil and botulinum toxin type-A, which have been reported to be beneficial if injected either alone or in combination with steroids.(10) Laser therapy for treating HTS is a relatively new concept started in the 1980s, being used initially to treat port-wine stains and remove decorative tattooing.(11, 12) Although its use is becoming more widespread for the reduction of established scars, its efficacy and mechanism of action remain to be established. Three main methodological variants of laser therapy have been developed over the years to treat specific aspects of established scars: Pulsed-dye lasers, Q-switched Nd:YAG lasers and fractional lasers.(13) Pulsed-dye laser therapy was used to reduce scar vascularity by inducing disruption of the targeted capillaries. This method was also reported to reduce itch.(14) Nd:YAG lasers emit light in the infra-red range, typically with a wavelength of 1064nm and have deeper tissue penetration.

Recently, twelve RCTs of laser therapy for the treatment of HTS, involving 592 patients, were considered in a systematic review.(15) Although 11 of these trials reported a positive effect of the therapy the review concluded there was insufficient evidence of the clinical effectiveness of laser therapy. This was largely due to variations in the laser therapy used, the scar assessment methods selected and inadequate study design. Currently there are 3 RCTs open in Canada and one in the US, this promising new therapy thus remains to have its clinical efficacy in scar management confirmed.

### Fractional carbon dioxide laser therapy

Fractionated CO_2_ laser therapy was introduced by Manstein et al in 2004 and essentially bridges the gap between the ablative and non-ablative laser techniques.(16) Ablative laser treatments work mainly on the epidermis and non-ablative treatments work solely on dermal collagen, fractional laser treatment works at both the epidermal and dermal layers of the skin making it suitable for treating several aspects of HTS. The CO_2_ lasers have a wavelength of 10,600nm which can be heavily absorbed by water. One study has suggested that a CO_2_ laser pulsed at less than 1ms, can limit the residual thermal damage to 100-150µm layer of skin by vaporizing tissue up to 20-30µm per pulse.(17, 18) It is thought that CO_2_ lasers allow immediate contraction of the ablative areas by denaturing the existing old collagen and stimulate new collagen formation.(18, 19) However, whether this is all that laser therapy does to achieve scar reduction seems unlikely and until we fully understand its mode of action this will limit approaches to improve its efficacy.

The Lumenis UltraPulse Encore model with DeepFX™ head piece is an advanced fractional CO_2_ laser system with three modes to deliver its energy: ActiveFX™, DeepFX™ and TotalFX™. The feature of DeepFX™ is that it focuses the laser’s energy into a 0.12mm spot size and allows for deep ablation of the tissue which can be useful for treating HTS resulting from deep partial thickness burn injuries. The principle of the Lumenis UltraPulse Encore DeepFX™ is in line with most of the CO_2_ lasers in that it introduces a beam of laser energy into the skin that generates an area of microscopic thermal injuries and allows the body to create a more rapid wound healing process and promote extracellular remodelling.(17, 20) It has been suggested that repeated treatments have continuous effects and facilitate scar tissues to remodel to a more normal and smooth appearance.(21) Because of the versatile and advanced features of this instrument, it will be used in this pilot study.

### Scar assessment tools

To assess scar development over time, the use of reliable scar assessment tools is crucial. Currently, subjective scar assessments such as Patient Observer Scar Assessment Scale (POSAS), Vancouver Scar Scale (VSS) and Brisbane Burn Scar Impact Profile (BBSIP) have been widely used in clinical practice. These assessments are in the form of questionnaires to reflect patients’ satisfaction as well as clinician’s perspectives toward the scars. However, these questionnaires only provide qualitive and/or semi-quantitative measures for post-burn scar assessment (22, 23). With the recent introduction of objective scar assessment tools, such as 3D-cameras (Eykona, Lifeviz and Vectra H1), DSMII and DSMIII ColoriMeter^®^, Cutometer^®^ and DermaScan^®^ high frequency ultrasound, accurate and reproducible evaluation of scars is made possible.(22) These quantitative tools measure different properties of the scar. For example, DermaScan^®^ measures scar thickness and density, DSMIII ColoriMeter^®^ quantifies colour, Cutometer^®^ assesses skin elasticity and the 3D-camera allows measuring the surface area and the volume of the scars. Contrary to subjective scar assessments, these quantitative assessment scores do not reflect patients’ satisfaction towards scars. With patients’ overall wellbeing in mind, to only report one area of the scar property may not represent the whole picture of the scar recovery. Therefore, a standardised scar assessment tool that incorporates objective scar assessment tools with the patient’s subjective satisfactory scoring systems would represent a more thorough and reliable assessment. In this study, a combination of assessment tools will be used to assess the clinical outcomes of the laser treatment.

### Cellular mechanisms

The cellular and molecular mechanisms behind laser therapy remains under researched. What is known so far is that dermal fibroblasts have been identified as the major player in skin wound healing (24) and it is likely that they will also be important in the response to laser therapy. One potential mechanism is the induction of cell senescence in response to the damage inflicted by the laser. Despite senescent cells playing a negative role in ageing (25) they also have positive effects. In recent years the beneficial role of cellular senescence in wound healing has been revealed. A study using a p16-3MR-transgenic mouse model in which senescent cells were deleted as they arise revealed slower wound healing. (26) The positive effects of senescent cells are likely mediated through their secretome, the senescence-associated secretory phenotype (SASP), which contains a range of pro-inflammatory cytokines, proteases and growth factors. The senescent cells appeared very early in response to a cutaneous injury, where they accelerate wound closure by inducing myofibroblast differentiation through the secretion of platelet-derived growth factor AA (PDGF-AA).(26) Another study provided evidence for the crosstalk between senescent fibroblasts and keratinocytes in human skin through a novel SASP factor contained in extracellular vehicles, namely miR-23a-3p which accelerates wound healing *in vitro*.(27) Given the central role of dermal fibroblasts and the beneficial role of senescent cells in promoting wound healing, it is likely the two cell types also participate in the tissue remodelling process post-laser therapy.

### Study objectives and aims

The study’s primary objective is to assess the kinetics of the response to ablative fractional CO_2_ laser therapy in HTS with respect to the cell types present in the treated skin. We will examine the presence of p16INK4A and γH2AX positive senescent cells, and CD90/Thy1^+^- and αSMA^+^-fibroblast subpopulations together with the combined subjective and objective scar assessments to measure the association between induction of senescent cells and scar reduction after laser treatment.

We suggest that improved understanding of the mechanisms that mediate scar reduction with laser treatment, will enable better design of laser treatment regimens, and thus benefit those living with HTS.

## Materials and methods

### Study design

SMOOTH (A prospective intra-patient **<u>S</u>**ingle-blinded randomised trial to examine the **<u>M</u>**echanistic basis of fracti**<u>O</u>**nal ablative carb**<u>O</u>**n dioxide laser **<u>T</u>**herapy in the treating adult burns and/or trauma patients with **<u>H</u>**ypertrophic scarring) is a multicentre, intra-patient, single-blinded randomised controlled longitudinal pilot observational study with parallel assignments, to measure the effects of ablative fractional CO_2_ laser on HTS in two anatomically comparable and independent scars per adult participant. The scars selected are randomly allocated to receive either ablative fractional CO_2_ laser therapy or standard care. An independent assessor will be blinded to the intervention and the control scar sites. The sample size will be a maximum of 60 patients, who will each have 3 laser treatments over a 12-month treatment period. After laser treatments, all patients will be followed up until 6-months after the 3^rd^ laser treatment. Biopsies will be taken at pre-treatment (visit 1), and 3-weeks (visit 2), 6-months (visit 4) and 12-months post 1^st^ laser treatment (visit 5); and subsequently assessed for the proportion of senescent cells and fibroblast subsets in the scars. Scar assessments will be performed at specified time points, including pre-laser treatment (visit 1), during laser treatment and post laser treatment (visit 3, 4, 5) to record the scar properties and the psychometric outcomes. Moreover, the study will also collect serum and plasma samples to assess the cellular and molecular biomarkers in response to laser therapy. **Fig 1** outlines the SPIRIT schedule of enrolment, intervention, and data collection. A diagrammatic overview of the SMOOTH trial study design is shown in **Fig 2**. SPIRIT reporting guidelines (28) were used throughout this study (**S1 File**).

**Fig 1.**
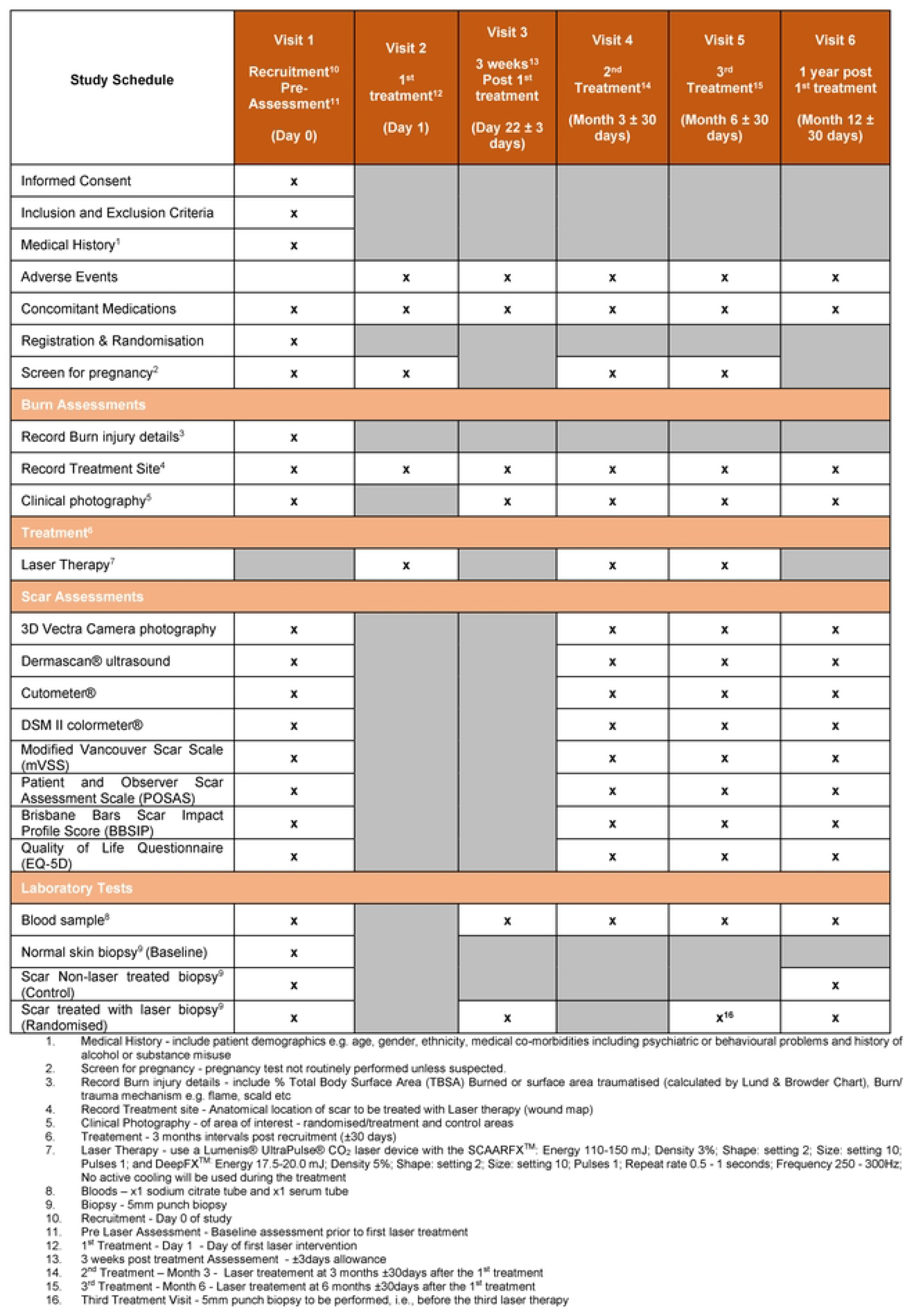
SPIRIT schedule of enrolment, interventions, and data collection.

**Fig 2.**
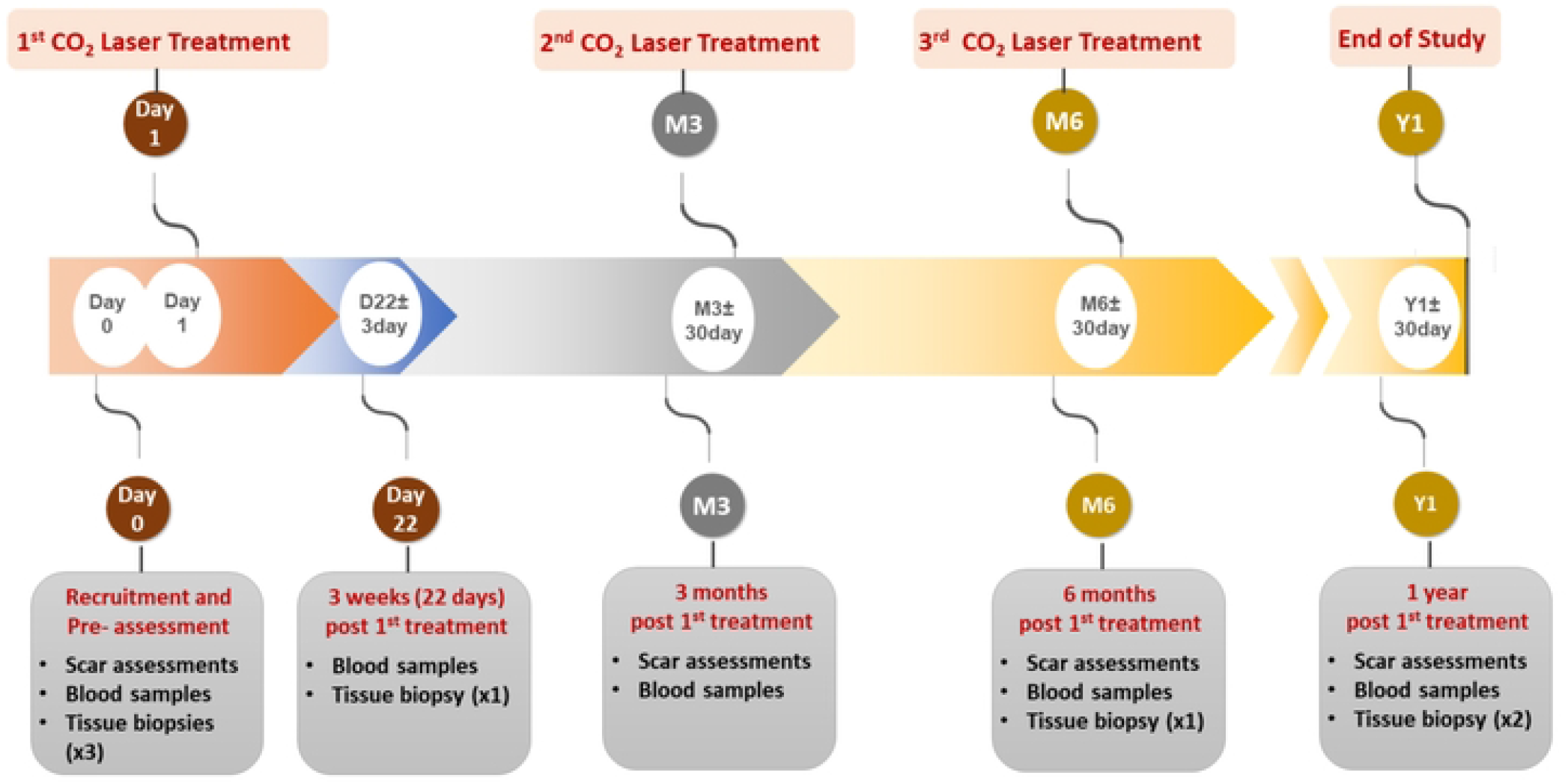
Diagrammatic overview of SMOOTH trial study design.

### Study setting

The study will be set in two UK-based hospitals: University Hospital Birmingham and The Welsh Centre for Burns and Plastic Surgery, Morriston Hospital, Swansea, UK.

### Study population

SMOOTH is a prospective study and following recommendations for pilot studies,(29) 30 patients or more are required to gain estimates of the parameters needed for sample size estimation. We have allowed for a 20% drop-out and possible loss to follow-up and therefore a total of 60 adult participants will be recruited to the study over a 2-year period. This will also allow the recruitment and retention rates to be estimated with 95% confidence interval maximum widths of 20% and 25% respectively. We aim to recruit both civilian and military veterans for this study. Civilian patients will be recruited from the Advanced Scar Management clinic at University Hospital Birmingham and The Welsh Centre for Burns and Plastic Surgery, Morriston Hospital, Swansea. Veterans will be recruited in collaboration with the CASEVAC Club volunteers. Participants will be identified by research clinicians or referred to the burns research team for screening by their primary clinician. Participants will be screened for study eligibility using the following recruitment criteria.

### Eligibility criteria

The main inclusion criteria are adult patients aged ≥16years with symptomatic HTS as a result of deep dermal or full thickness burns or trauma that were sustained more than 12 months previously. Patients should have had no previous laser therapy treatment to the study site and the treatment area must be ≥25cm^2^ of confluent scarring with a comparable control scar on the limb or trunk.

Patients with concurrent use of pressure garments, emollient application and scar massage will also be included in the study. However, if the patient has recent or concurrent invasive scar treatments, including intralesional pharmaceuticals, micro-needling or other laser modalities (e.g. pulse-dye) on the study site, they will be excluded. Patients with known allergy or contraindication to eutectic mixture of local anaesthesia (ELMA) cream (lidocaine 2.5% and prilocaine 2.5%), components of moisturising cream (benzalkonium chloride 0.1%; chlorhexidine dihydrochloride 0.1%; liquid paraffin 2.5%; isopropyl myristate 2.5%) or ointment (white soft paraffin liquid paraffin %w/w 50/50); and patients with Fitzpatrick skin type of 5-6 due to nature of the skin will also be excluded from the study. The main recruitment criteria can be seen in **Fig 3**.

**Fig 3.**
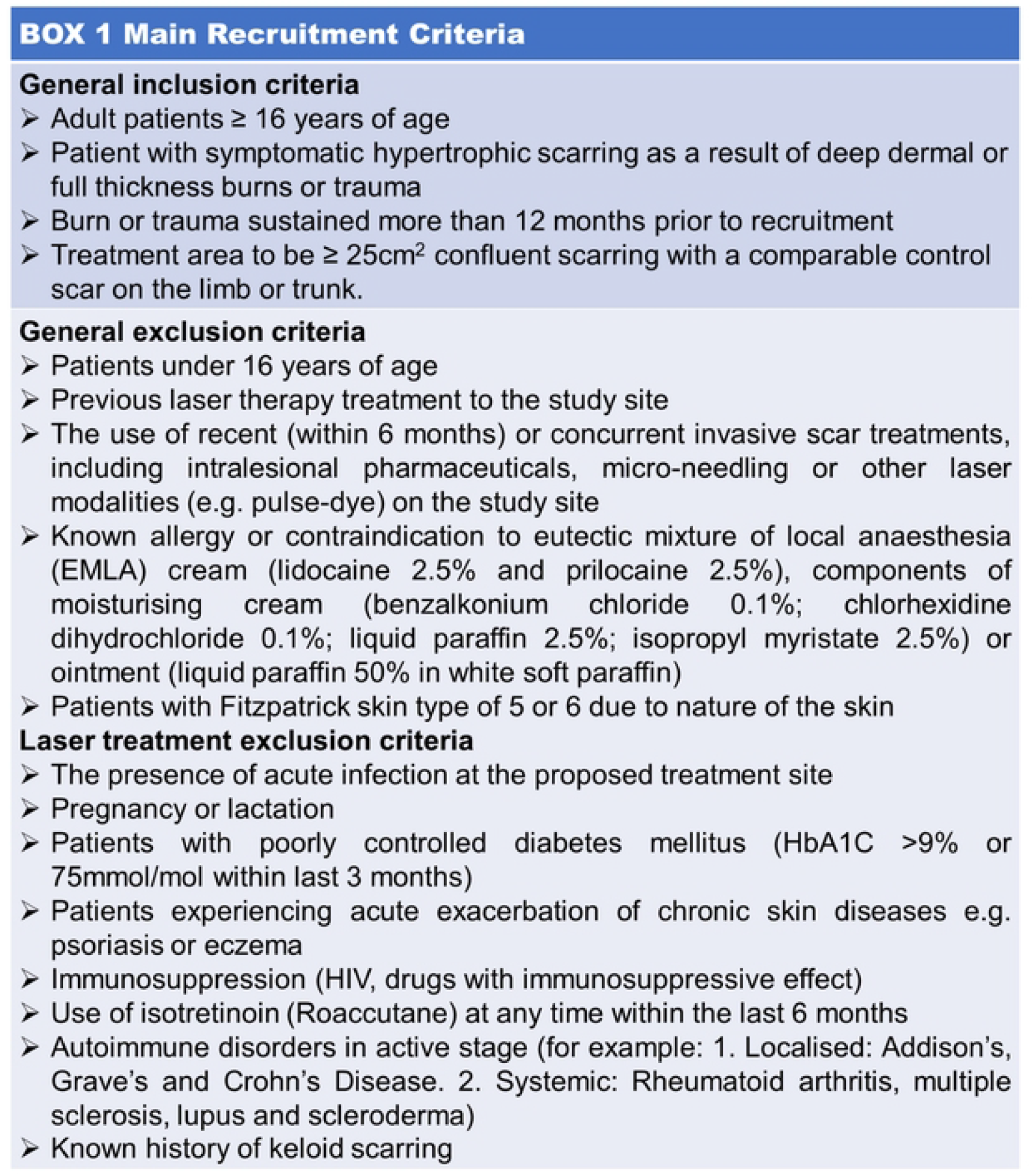
Main recruitment criteria.

### Identifying participants and enrolment

Patients will be identified through the Advanced scar management clinic, burns and scar therapist outpatient clinics at QEHB and consultants/therapy led clinics at Morriston Hospital, Swansea. Patients who are deemed suitable will be approached in person by a member of the research team or via telephone during which a brief explanation of the trial will be given. If the patient agreed, a Patient Information Leaflet (PIL) will be sent by mail or given to the patient (**S2 File**).

### Consent and withdrawal from study

Written informed consent will be obtained from all participants before the start of the study. The research team will assess patient’s eligibility for each treatment intervention, i.e., first, second and third laser treatment, and re-confirm patient’s consent to continue with the treatment at each time point.

Participants may withdraw or be withdrawn from the study at any time if an incident occurs that renders the participant unable to continue with the study. For example: (1) patients unable to complete all laser therapy sessions as planned; (2) adverse reaction to laser therapy and (3) pregnancy. The reason for discontinuation will be collected and recorded in the electronic case report form. Any data collected at the time of withdrawal may still be included in the data analysis, unless the participant specifically withdraws their consent. Participants will be asked to clarify this at the point of withdrawal.

### Randomisation and blinding

Each participant will have their own control scar in an anatomically comparable site either on the trunk or limbs. Allocation of scar treatment will be performed using a computer-based randomisation system developed at the University Hospitals Birmingham NHS Foundation Trust (UHBFT). Two comparable anatomically scarred areas ≥25cm^2^ on trunk, arms or legs will be identified on the same participant and will be described as Scar-A and Scar-B on the body map. The participants’ identified scar sites, A and B, will be randomised on a 1:1 basis to either the standard of care or laser treatment as shown in **Fig 4**.

**Fig 4.**
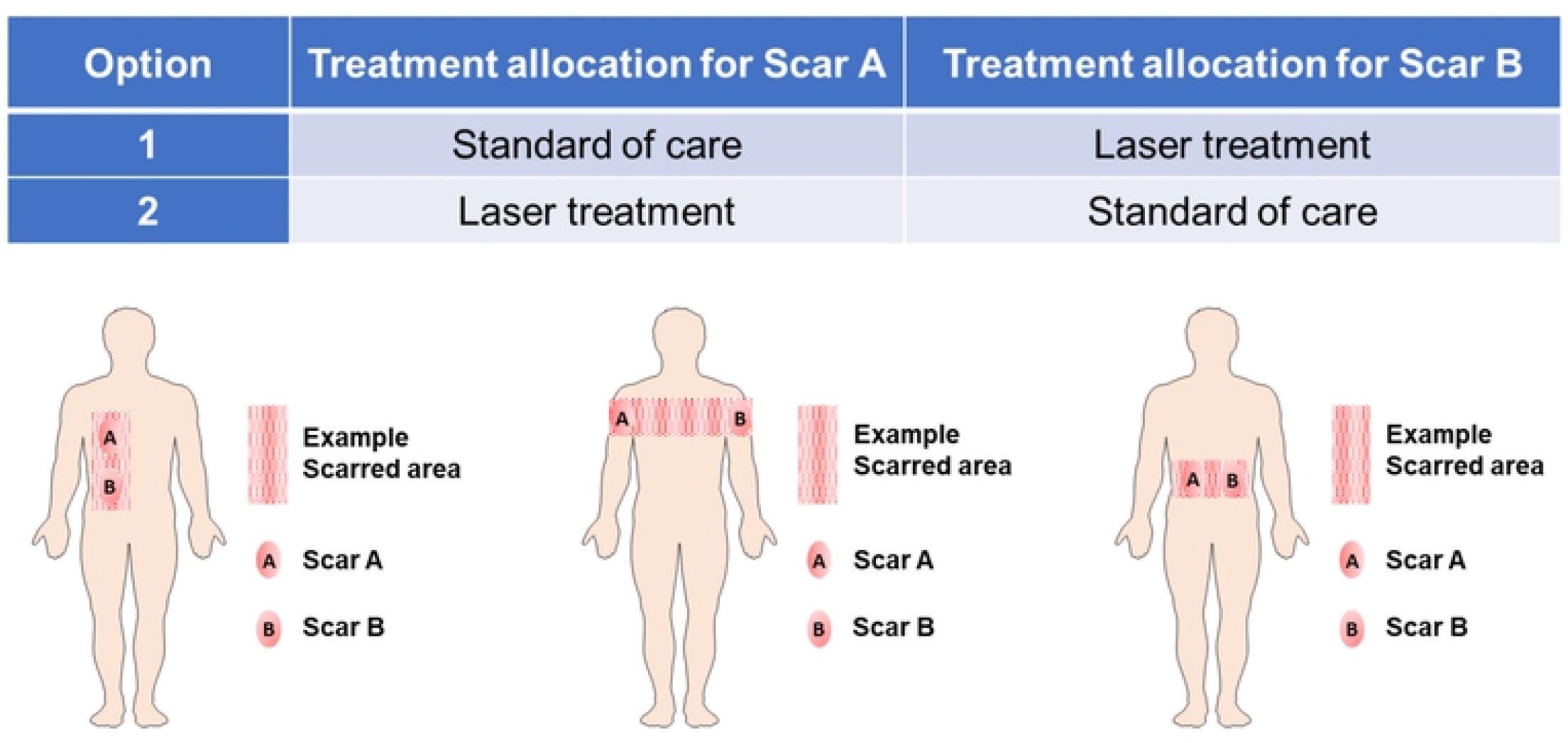
Options of assignment of study areas.

### Study schematic

This study is a prospective intra-patient single-blinded randomised controlled pilot study designed to examine the mechanistic basis of ablative fractional CO_2_ laser therapy in hypertrophic scarring **(Fig 3)**. Patients fulfilling the recruitment criteria will be enrolled into the study. During the pre-assessment visit D0 (visit 1), serum, plasma and three biopsies from one normal control area, and two scar sites (control and intervention) will be obtained. Scar assessments including modified VSS (mVSS), POSAS, BBSIP and European quality of life five dimension (EQ-5D) and objective scar assessments including 3D-Vectra H1 Photography, DermaScan^®^, Cutometer^®^ and DSMIII ColoriMeter^®^ will also be performed and recorded. The scars will then be randomised after pre-assessment visit using a computer-based randomisation system developed at the UHBFT.

During the treatment and assessment visits, patients will receive three laser treatments and time points will occur at 3 months intervals post-recruitment D1 (visit 2), M3 (±30 days; visit 4) and M6 (±30 days; visit 5). Prior to each laser treatment schedules, there will be assessment visits where serum, plasma and biopsy samples are collected. The serum and plasma will be collected at all three assessment visits (visit 3, 4 & 5). The biopsies from the treatment site of the scar will be collected at 1^st^ and 3^rd^ assessment visits (visit 3 & 5). Scar assessments will also be performed and recorded during 2^nd^ and 3^rd^ assessment visits (visit 4 & 5). After 3 sessions of laser treatment schedule, there will be a follow-up visit 1 year post the 1^st^ laser treatment (visit 6). During this visit, scar assessments will be performed; serum, plasma and biopsies from control and treatment sites will be collected **(Fig 5)**.

**Fig 5.**
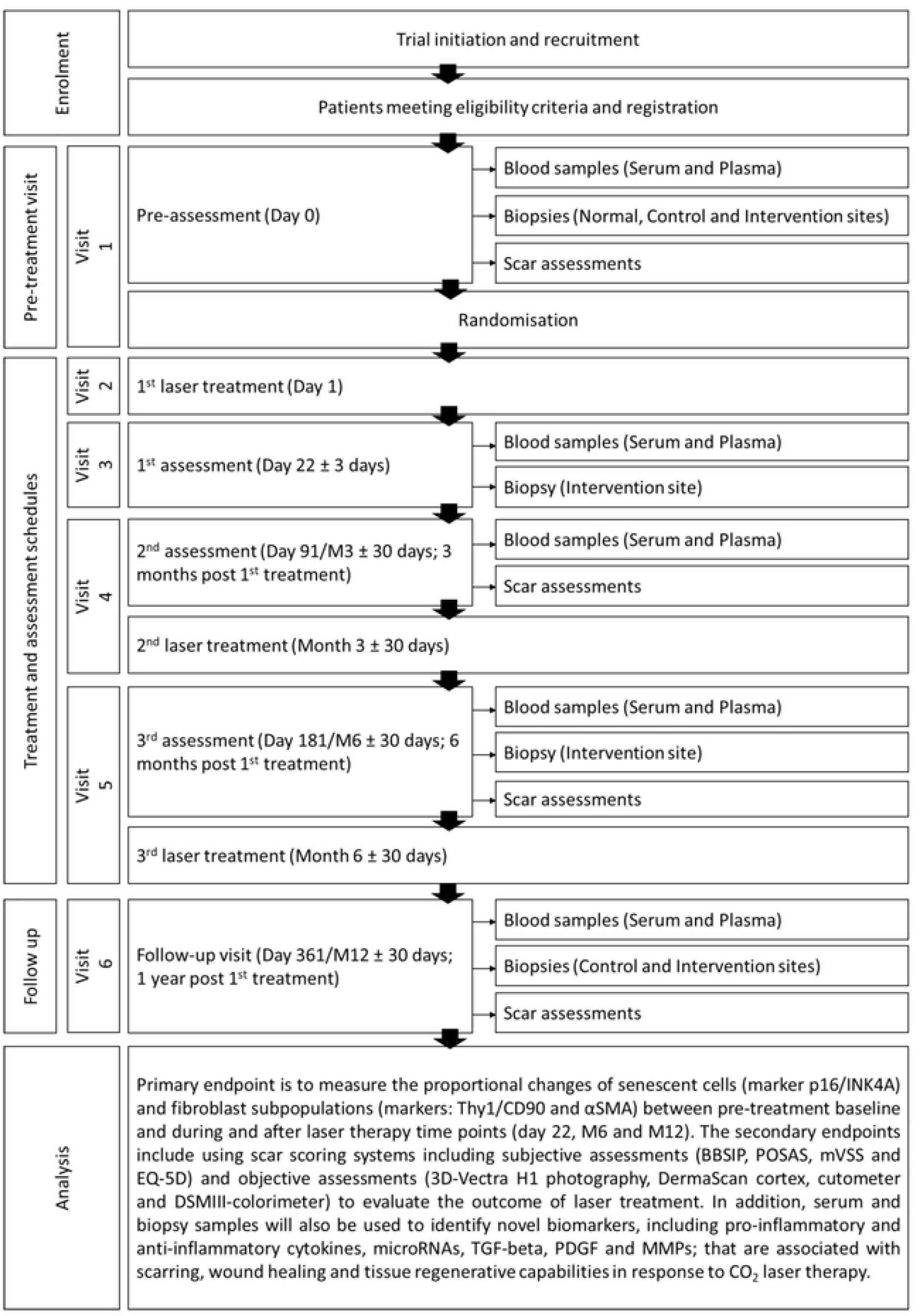
Overview of SMOOTH study schematic.

### Biological measurements

5mm skin punch biopsies will be collected under local anaesthesia from each patient. Three different types of biopsies will be collected at various time points: (1) Normal skin biopsy: pre-assessment visit (visit 1); (2) Control, scar without laser treatment: pre-assessment and follow-up visit (visit 1 & 6); and (3) Intervention, scar with laser treatment: pre-, 1^st^-, 3^rd^-assessments and follow-up visit (visit 1, 3, 5 & 6). All biopsies collected from patients will be processed immediately and bisected with one half transferred to a fixative solution (10% neutral buffered formalin) for downstream histological and immunohistochemical assessments. From histological assessment, dermal and epidermal thickness, collagen structure and orientation, elastin density and structure will be recorded, and from immunohistochemical assessments, the proportion of p16INK4A^+ve^ and γH2AX^+ve^ senescent cells, and fibroblast subpopulations will be measured to investigate variables of scar behaviour in response to laser therapy. The other half of the biopsy will be frozen in CryoStor^®^ cell cryopreservation media (Merck Life Science UK Limited, Dorset, UK) and stored at −80°C for future analysis using single cell RNA sequencing (scRNAseq) to delineate the full profile of skin cells present during scar reduction process in response to laser therapy.

Serum and plasma will be collected on pre-, 1^st^-, 2^nd^- and 3^rd^-assessment visits (visit 1, 3, 4 & 5) and follow-up visit (visit 6). Full white blood cell count will be recorded using Sysmex XN-1000 haematology analyser (Sysmex UK, Milton Keynes, UK). Serum cytokine patterns will be determined using Luminex Assay and a range of pro- and anti-inflammatory cytokines (IL1β, IL4, IL6, IL8, IL10, IL13, IL1Rα) measured. Other factors measured will include those known to influence scarring, such as TGFβ, decorin, PDGF-AA and adiponectin. Double-spun plasma will also be prepared(30) for future analysis of the role of extracellular vesicles and their cargo in scar reduction.

### Scar assessments

Scar assessments for both treated and control scars will be performed and recorded, prior to randomisation and before each laser treatment (visit 1, 4, 5 &6). The control scar will be treated as per standard of care, however if the study participant desires laser therapy to the area, this would be done at the 6-month follow-up after study completion. All study visits are aligned with standard of care treatments and no additional follow-up visits would be expected.

To associate the findings of the biological measurements with the clinical outcomes, the study will incorporate qualitative and quantitative scar assessments to record scar properties and psychometric outcomes to reflect the clinical effects after laser therapy. Patient reported outcome data will be recorded using a series of patient reported outcome measures (PROMs) and scar assessments. Scars will be assessed using tools including mVSS, POSAS and BBSIP. Scars will also be objectively assessed using 3D-Vectra Photography for scar site identification, DermaScan^®^ for scar thickness and density, Cutometer^®^ for elasticity and colour using DSMIII Colorimeter^®^. As part of the evaluation of the impact of laser treatment on patients’ QoL and the patient reported outcome strategy,(31) a PROMS validation study using Rasch analysis will be carried out to evaluate if the PROMs are useful measurement tools for evaluating the impact of laser therapy on scar tissue and QoL.(32)

Scar assessments tools including POSAS and mVSS will be performed by an independent and experienced assessor in scar management and treatment. The health care professional or scar assessor, in addition to the data analysts, will be blinded to the anatomical site (treated scar) receiving laser treatment compared to the non-laser treated scar (control scar). Scar assessment questionnaires, including BBSIP, POSAS, as well as QoL questionnaire EQ-5D will be completed unblinded by participants.

All study visits are aligned with standard of care treatments and no additional follow-up visits would be expected. The full trial schema can be seen in **Fig 5**.

### Trial intervention

The treatment area is ≥25cm^2^ confluent scarring with an anatomically comparable control scar on the limb or trunk.

### Control Area (Standard of Care)

Patients with HTS usually undergo massage therapy, pressure garment application and/or steroid or fluorouracil injection as part of standard practice. The control scar, if deemed of inferior quality to the laser treated area, will be treated as part of the patient routine ongoing management of their scars after the trial is completed.

### Treatment Area (Intervention)

The laser treatment will be performed under local anaesthetic and will be conducted by a suitably trained medically qualified doctor. The treatment will use a Lumenis^®^ UltraPulse^®^ CO_2_ laser device with the DeepFX™ and/or SCAARFX™ headpiece depending on scar thickness assessed by ultrasound prior to treatment. The proposed treatment area will be marked and photo documented. The treatment will include a single pass of the chosen treatment site with the following settings: SCAARFX™: Energy 110-150mJ; Density 3%; Shape: setting 2; Size: setting 10; Pulses 1; and DeepFX^TM:^ Energy 17.5-20.0mJ; Density 5%; Shape: setting 2; Size: setting 10; Pulses 1. Repeat rate 0.5 seconds; Frequency 300Hz; No active cooling will be used during the treatment.

Laser treatment during the trial will only be carried out on the scar allocated to receive laser treatment. If laser treatment will be done on additional areas, then the scar allocated to receive laser treatment and the same additional site and size will need to be treated based on the time points mentioned in this protocol throughout the study period. This is to ensure that the patient receives the same dose of laser treatment for consistency of assessment throughout the study.

### Concomitant therapy

Control and laser treated (randomised) scars, when healed, will be treated as per standard of care which would include silicone, massage and pressure therapy, when applicable.

### Study objectives

The study’s primary objective is to assess the kinetics of the response to ablative fractional CO_2_ laser therapy in HTSs with respect to the cell types present in the treated skin. We will examine the presence of p16INK4A^+ve^ and γH2AX^+ve^ senescent cells, and CD90/Thy1^+^- and αSMA^+^-fibroblast subpopulations together with the combined subjective and objective scar assessments to measure the association between induction of senescent cells and scar reduction after laser treatment.

### Outcome measures

During the study, parameters including histological examination of scars following laser therapy will be measured. In addition, treatment outcomes, including both subjective scar assessments (POSAS and mVSS) and objective scar assessments (scar thickness, density, pliability, and colour) will be recorded. Patient reported outcomes will also be measured using BBSIP. Outcome data will be collected at pre-laser, 3 weeks-, 6 months- and 1 year-post the 1^st^ laser treatment.

Our primary outcome measure is to assess the proportional changes of senescent cells in skin and the changes of sub-populations of fibroblasts using cell markers αSMA and CD90/Thy1 in HTS after laser therapy via immunohistochemical staining. Secondary outcome measures comprise a range of scar assessment tools, including mVSS, POSAS, BBSIP and EQ-5D generic health status assessments. Scars will also be assessed through histological examinations to assess dermal and epidermal thickness, collagen structure and orientation, elastin density and structure following treatments.

Exploratory measures for novel markers associated with scarring and tissue regenerative capabilities such as extracellular vesicle associated microRNAs, cytokines, TGFβ, decorin, PDGF-AA and adiponectin will also be assessed. As part of the patient reported outcome strategy, we will evaluate the extent to which the PROMs are sound psychometric measures of improvements in scarring using Rasch analysis, and the impact on participant’s QoL as a result of laser treatment. So as not to miss any novel cells that may be induced, we will also use scRNAseq to fully characterise cells present in the laser treated tissue. This technique has been used previously to identify the role of Engrailed-expressing fibroblasts in the scarring process in mice.(33) This technique will also allow us to determine the transcriptomic response to therapy to further define the mediators of the beneficial effects of laser therapy.

### Data Management

#### Data analysis plan

Following recommendations for pilot studies, 30 patients or more are typically required to gain estimates of the parameters needed for sample size estimation. No formal sample size calculation has been performed as a result. We have allowed for a 20% drop-out and possible loss to follow-up. We therefore aim to recruit 60 patients in total. This will also allow the recruitment and retention rates to be estimated with 95% confidence interval maximum widths of 20% and 25% respectively.

All analysis will be based on the intention to treatment principle. The primary comparison groups will be the scar sections randomised to standard of care (control group) versus those randomised to treatment with laser therapy (experimental group). The data analysis for this pilot study will be descriptive and mainly focus on confidence interval estimation, with no formal hypothesis testing performed. Dichotomous feasibility measures, such as the recruitment and retention rates, including completeness of data will be reported as numbers and percentages. These values will be summarised across participants or treatment groups as appropriate.

Analysis methods will be chosen according to the data type: 1. Continuous endpoints (e.g. mVSS total score): These data will be summarised using means and standard deviations, with differences in means with 95% confidence intervals reported. Longitudinal plots of the data over time will also be constructed for visual presentation of the data. 2. Categorical (dichotomous) endpoints (e.g rates of improvement in scar domains): the number and percentages of participants/scars experiencing the event will be summarised across and between groups. For exploratory outcome data about the biomarkers of the scar response to laser therapy, the various cellular and molecular variables will be examined for relationships to scarring using logistic regression.

#### Data management

The data collection tool for this study will be both paper and electronic CRFs. The data collection may be conducted by the CRN nurses, the PIs, the health care professionals or the research fellows involved in the study. Collected data will be entered electronically onto the online trial database, REDCap Cloud. The database REDCap cloud is run by the Birmingham Surgical Trials Consortium (BiSTC), University of Birmingham, under licence from Vanderbilt University. Primary analysis for the trial will occur once all participants have completed the 12-month assessment and corresponding outcome data has been entered onto the trial database, validated as being ready for analysis, and the database locked. This analysis will include data items up to and including the 12-month assessment.

#### Data monitoring

All data collected will be recorded and stored in the trial site file, including the original signed informed consent form (**S3 File**), will be stored in the recruitment sites for quality control purposes. Patient’s confidentiality will be maintained and follow the General Data Protection Regulation guidelines at all times and data will not be disclosed without written consent. Principal investigator will be responsible for the secure storage of all trial related documentation. All documentation including copies of protocols, patient’s information leaflets, GP letters, consent forms, and CRFs will be held securely in accordance with current ICH GCP guidelines for a minimum of fifteen years. The records will be available for review by governing bodies upon request following notice. All trial documents will be archived in accordance with the UHBFT Archiving Procedures.

### Patient and public involvement

UHB’s Accident, Burns and Critical Care (ABC) PPI group have been involved in the development stage of the proposed study up to the finalisation of the study protocol and study documents.

### Ethics and dissemination

The SMOOTH Study is conducted according to the Declaration of Helsinki (2008). It received favourable opinion on from the North Scotland Research Ethics Committee (REC reference: 19/NS/0125). Consequently, on 22^nd^ September 2022, the Health Research Authority (HRA) and Health and Care Research Wales (HCRW) issued the approval. This article refers to protocol v.7.0 dated 22^nd^ September 2022 (**S4 File**). Results will be disseminated via peer-reviewed publication and presentations at national and international conferences.

### Status and timeline of the study

The SMOOTH study is an ongoing trial with an estimated finishing time by the end of August 2023.

### Trial registration

ClinicalTrials.gov: NCT04736251. Trial registration dataset: **S1 Table**.

## Discussion

Hypertrophic scarring is one of the major concerns after burn injury, with detrimental physical and psychological consequences. Despite laser therapy has been used for treating HTS for some time with various positive outcomes, the mechanism of actions remains not fully understood. In this study, we used a longitudinal study design to enable us delineating the cellular kinetics within the hypertrophic scars after laser treatment. In order to help us to limit the variations due to physiological differences from different anatomical sites, we applied an intra-participant randomisation type 1:1 control and treatment comparable area in the same participant. Furthermore, to represent a more thorough and reliable overall clinical outcome, we incorporate both objective scar assessment tools with subjective scoring systems to evaluate treatment outcomes. With this study protocol, we believe it will provide us an improved understanding of mechanisms mediating scar reduction with laser treatment, which will enable better design of laser treatment regimens, and thus benefit those living with HTS.

## Data Availability

No datasets were generated or analysed in the current study protocol. Deidentified research data will be disseminated via peer-reviewed publication and presentations at national and international conferences once the study is completed.

## Roles and responsibilities

### Sponsor contact information

University Hospital Birmingham Foundation Trust. Mindelsohn Way, Edgbaston, Birmingham, B15 2WB.

### Funder information

This work is supported by the Chancellor using LIBOR funds via a grant obtained by The Scar Free Foundation.

### Authors’ contributions

NSM is the chief investigator and was responsible for the design and development of the SMOOTH study. JML, YW and JY are principal investigators that assisted in protocol development and/or design. JB, AS, YYC, KMP and TH assisted in protocol design. YYC, KMP, RI, TH, EA, AA and MV commented on the protocol and are involved in the acquisition of data. YYC, KMP and RI wrote the manuscript and all authors revised and approved the final version.

## Acknowledgement

This project is funded by the Scar Free Foundation. The authors would like to thank the SFF, University Hospital Birmingham Foundation Trust, National Institute for Health and Care Research, the UHB’s Accident, Burns and Critical Care (ABC) PPI group and the patients involved in this study.

## Supporting information

**S1 File. SPIRIT checklist**

**S2 File. Modelled SMOOTH patient information leaflet v7.0**

**S3 File. Modelled SMOOTH informed consent form**

**S4 File. SMOOTH protocol version 7**

**S1 Table. Trial registration dataset**

